# Financial Incentives for Quitting Smoking in Pregnancy: Are they cost-effective?

**DOI:** 10.1101/2022.06.21.22276693

**Authors:** Nicola McMeekin, Lesley Sinclair, Lyn Robinson-Smith, Alex Mitchell, Linda Bauld, David M Tappin, Kathleen A Boyd

## Abstract

**Aims:** To evaluate whether adding financial incentives to usual care is cost-effective in encouraging pregnant women to quit tobacco smoking, compared to usual care alone.

**Design:** Cost-effectiveness analysis (CEA) and cost-utility analysis (CUA) from a healthcare provider’s perspective, embedded in the Smoking Cessation in Pregnancy Incentives Trial (CPIT III). Long-term analyses were conducted from the same perspective, using an existing Markov model over a lifetime horizon.

**Setting:** Seven maternity smoking cessation sites in Scotland, England and Northern Ireland in the United Kingdom.

**Participants:** In the short-term analysis CPIT III participants were assessed: women 16 years or older, self-reporting as smokers, less than 24 weeks pregnant and English speaking (n=944). The same population was used for the lifetime analysis, plus their infants.

**Measurements:** Costs include financial incentive vouchers and postage, cessation support and nicotine replacement therapy and neonatal stays. The outcome measure was biochemically verified quit rate for the CEA and quality adjusted life-years (QALY) for CUA. Costs are presented in 2020 GBP sterling (£).

Data for the lifetime analysis came from the trial and was combined with data from published literature embedded in the model, reporting incremental cost per quitter and QALY. A 3.5% discount rate was applied.

**Findings:** The short-term incremental cost per quitter was £4,400, uncertainty in QALY gains resulted in a cost per QALY of £150,000. Results of sensitivity analyses confirm these results. The long-term analysis combined costs and outcomes for mother and infants, results show a cost saving of £37 (-£35 to £106) and increase in QALYs of 0.171 (0.124 to 0.229). These findings indicate that, over a lifetime, financial incentives are cost saving and improve health outcomes.

**Conclusions:** Offering up to £400 financial incentives, in addition to usual care, to support pregnant women to stop smoking is cost-effective over a lifetime for mother and infants.

**Ethics:** Ethics approval received from NHS West of Scotland Research Ethics Committee-2, August 2017.

**Registration details:** Trial registration number: ISRCTN15236311, date registered 09/10/2017 https://doi.org/10.1186/ISRCTN15236311

## INTRODUCTION

Tobacco smoking is the principle cause of preventable deaths globally; linked to eight million deaths annually worldwide and over 91,000 in the United Kingdom (UK)(1). Tobacco smoking prevalence during pregnancy is 1.7% globally(2), however in the UK it is higher. In 2020/21 it was 9.6% in England(3) and 13% in Scotland(4). Smoking during pregnancy is linked to low birth weight and increased risk of premature birth(5). After pregnancy passive smoking is linked to an increased risk of sudden infant death syndrome, lower respiratory diseases, asthma and impaired lung function in infants(5). Children living in households with smokers are also 90% more likely to take up smoking than children living in non-smoking households(6). In addition to the burden on health to mother and infant, the economic burden of smoking during pregnancy is estimated to be over £23 million annually in UK(7). As 80% of women in UK have a baby(8), pregnancy poses a good opportunity to quit smoking, improving the health of the mother and infant, and reducing pressure on healthcare budgets.

All pregnant women in the UK are offered National Health Service (NHS) support and nicotine replacement therapy (NRT) to stop smoking, but few use this service and set a quit date (11%) and even fewer (3.5%) remain abstinent four weeks after their quit date(9). Financial incentives have been shown to be effective in supporting women to quit tobacco during pregnancy, a recent Cochrane review combined results of nine trials to estimate a relative risk of 2.38 (95% confidence interval (95%CI) 1.54 – 3.69), favourable to financial incentives(10). One of these nine trials was a single-site phase II trial in Scotland, Smoking Cessation in Pregnancy Incentives Trial II (CPIT II), which found offering a maximum of £400 financial incentives resulted in higher quit rates, this research was carried out by the CPIT trial team(11). CPIT II estimated a lifetime incremental cost per QALY of less than £500 which is considered highly cost-effective when compared to the National Institute of Health and Care Excellence (NICE) willingness-to-pay threshold of £20,000(12, 13). Following the CPIT II trial a financial incentive scheme was introduced in NHS Greater Glasgow and Clyde (NHSGG&C), this was found to be effective in improving quit rates at four and twelve weeks post-quit and the incremental cost per quitter was less than £550 for both four- and 12-weeks post quit date(14). Whilst this is encouraging, additional evidence is needed from multi-site research, with a longer follow-up period providing information on relapse post birth(15).

More recently, CPIT III evaluated the effectiveness of offering up to £400 financial incentives to support pregnant women to stop smoking(16). This paper reports the economic evaluation of CPIT III, exploring whether financial incentives are cost-effective in encouraging pregnant women to stop smoking.

## METHODS

### CPIT III overview

CPIT III was a pragmatic, multi-centre, randomised controlled trial which assessed the effectiveness and cost-effectiveness of offering financial incentives in addition to usual care, compared to usual care only, to improve smoking quit rate in pregnant women. Participant inclusion criteria included pregnant women self-reporting as smokers, 16 years or older, less than 24 weeks pregnant and English speaking. Participants were recruited from seven sites across England, Scotland and Northern Ireland between February 2018 and April 2020.

The trial primary outcome was quit rate at late-pregnancy (34-38 weeks gestation) with those self-reporting as quit confirmed as abstinent by biochemical verification. Participants with missing primary outcome data were assumed to be smokers, as per best practice(17). Participants had further follow-up up to six-months post-partum to establish biochemically verified sustained quit rate.

Current Controlled Trials ISRCTN15236311 date registered 09/10/2017 https://doi.org/10.1186/ISRCTN15236311

### Economic evaluation

Two time horizons were considered; short-term within trial and lifetime, both taking an NHS and personal social services perspective. Cost-effectiveness was reported as cost per quitter and cost per quality adjusted life-year (QALYs).

This research follows best practice for methods (18, 19) and reporting of economic evaluations(20). The health economics analysis plan is available elsewhere(21).

### Within-trial analysis

The population for the within-trial analysis was that of the CPIT III trial. The time horizon for this analysis was from recruitment into the trial to birth, as this was less than one year discounting was not applied to costs or outcomes.

#### Treatment arms

The intervention arm consisted of financial incentives worth up to £400 in shopping vouchers plus usual care. Participants received a £50 voucher for engaging with stop smoking services (SSS) and setting a quit date. Participants who were carbon-monoxide (CO) verified as quit at 4- and 12-weeks post quit date received £50 and £100 vouchers, if they met the criteria for the previous voucher. Participants received a final £200 voucher if CO verified quit at late-pregnancy, participants could still receive this voucher if they had not met criteria for previous shopping vouchers. Due to the COVID-19 pandemic an amendment was made for participants reporting smoking status after 16th March 2020: self-reporting for quit at 4- and 12-weeks, and participants self-reporting as quit at late-pregnancy received the final voucher if they provided a saliva sample. The control arm was usual care only.

Service delivery varied between sites but usual care was typically SSS plus NRT for 12 weeks.

#### Resource use

Resource use categories include issued vouchers, postage for vouchers, SSS, NRT and neonatal costs.

The trial data management system recorded when vouchers were sent to participants and a standard postal charge was applied to each voucher. During the trial 87 vouchers were resent, 59 of which required a postage charge, this charge was also captured.

Individual participant details of SSS received and NRT prescribed were collected for five sites using NHS routinely collected data and bespoke Excel spreadsheets. For the remaining two sites, site-specific typical SSS and NRT use for an individual (established using expert opinion) was applied to participants reporting NRT use in the trial database.

Neonatal care stays were not collected during the trial so prematurity at birth was used as a proxy. Prematurity was classified by severity as defined by the World Health Organisation as follows: ‘extremely preterm’ (< 28 weeks gestation), ‘very preterm’ (28-32 weeks gestation) and ‘moderate to late preterm’ (32-37 weeks gestation)(22). Length of stay was applied to each class of premature birth: 93 days for extremely preterm, 44 days for very preterm and 13 days for moderate to late preterm(7).

#### Unit Costs

Unit costs were obtained from routine sources (Table 1) including British National Formulary (23), Personal Social Service Research Unit (24), NHS National Cost Collection (25) and the CPIT trial team. Costs are reported for the price year 2020 and expressed in pound sterling (GBP£).

**Table 1:** Unit costs.

| **Variable** | **Unit cost** | **Source** |
| --- | --- | --- |
| Voucher engagement and setting quit date | £50 | CPIT III Trial |
| Voucher 4-week CO verified quit | £50 |  |
| Voucher 12-week CO verified quit | £100 |  |
| Voucher late-pregnancy CO verified quit | £200 |  |
| Voucher postage | £2.92 per voucher |  |
| Resent voucher postage | £2.05 per voucher |  |
| Sensitivity analysis postage | £6 per voucher |  |
| Smokefree advisor/midwife per hour (Grade5/6 depending on site) | £39/£49 | PSSRU 2019/2020 |
| Neonatal costs per day (moderately pre-term) | £536.45 | NHS National Cost Collection version 2 2019/2020 Healthcare resource group<br>XA05Z Neonatal Critical Care, Normal Care |
| Neonatal costs per day (very pre-term) | £709.16 | NHS National Cost Collection version 2 2019/2020 Healthcare resource group<br>XA03Z Neonatal Critical Care, Special Care |
| Neonatal costs per day (extremely pre-term) | £1707.5 | NHS National Cost Collection version 2 2019/2020 Healthcare resource group<br>XA01Z Neonatal Critical Care, Intensive Care |

Unit costs were combined with resource use data to estimate a mean cost per participant in each trial arm. Generalised linear model (GLM) regression analysis was conducted to estimate the cost difference between arms, adjusting for site, baseline age, number of years of smoking and primary outcome collection pre- or post-16 March 2020(26).

#### Outcomes

Two outcomes were used, late-pregnancy quit rate and QALYs. The timing of late-pregnancy was used as a proxy for birth due to the difficulties of collecting data at birth. A QALY combines health-related quality of life and quantity (length) of life. Quality of life was measured using the EQ-5D-5L questionnaire(27), completed by participants at baseline, late-pregnancy and up to six months post-partum. Responses were converted to health utilities using the UK value set (28) and crosswalk mapping as recommended by NICE(29, 30). Quantity of life was measured by the length of time a participant remained in the trial. A standard area under the curve approach was used to calculate QALYs, with changes in utilities between follow-up points treated as linear(31, 32). GLM regression analysis was conducted to estimate the difference between arms, adjusting for site, baseline age and utilities, gestational age at booking and primary outcome collection pre- or post-16 March 2020(31).

#### Analysis of costs and effects

Mean costs and outcomes are presented with standard errors (SE) for each arm and differences between arms are presented with 95%CI. Cost per late-pregnancy quitter and cost per QALY incremental cost-effectiveness ratios (ICERs) are presented and the latter is compared to the UK NICE willingness-to-pay threshold of £20,000 to assess cost-effectiveness(13).

### Uncertainty

Uncertainty in the results was explored with non-parametric bootstrapping using 1,000 iterations(33, 34). The bootstrapped results were plotted on a cost-effectiveness plane. A cost-effectiveness acceptability curve (CEAC) for the QALY outcome is presented with varying willingness-to-pay thresholds to explore cost-effectiveness at different thresholds.

### Missing data

Missing data was assessed as missing at random and multiple imputation using chained equations was used to replace missing data at a disaggregated level, following best practice recommendations(33).

### Sensitivity analyses

Nine sensitivity analyses were conducted to explore the effects on the results of altering inputs, these were: 1) Including miscarriage as a covariate in the GLM regression; 2) using self-reported smoking status at late-pregnancy; 3) adjusting for possible gaming (where participants self-report quit, are biochemically verified quit yet a residual blood sample fails to confirm quit status), based on evidence from trial (incentives 2/18 11%, control 2/10 20%); 4) adjusting for possible gaming using incentives arm only (conservative estimate); 5) 6-month post-partum quit rate; 6) 6-month post-partum QALYs (from EQ-5D responses); 7) complete case analysis; 8) including postage per voucher at £6, and 9) including postage per voucher at £0. The latter two analyses were conducted to explore potential implementation scenarios.

Analysis was undertaken using Stata 17 (StataCorp. 2021. Stata Statistical Software: Release 17. College Station, TX: StataCorp LLC.).

### Lifetime analysis

In the short term we would not expect quit rates to translate into immediate health gains so long-term analysis is needed to reflect the impacts of improved quit rates. The lifetime analysis was conducted to capture all relevant costs and benefits of these impacts(18). The lifetime analysis utilised a published model developed for assessing the cost-effectiveness of interventions for smoking cessation in pregnant women(7). The discount rate used was 3.5% for costs and QALYs as recommended by NICE(13).

#### Model structure

The Economics of Smoking in Pregnancy (ESIP) model combines decision trees and Markov models to estimate an incremental cost per QALY for mother and infant over a lifetime (up to 100 years old) (Figure 1). Further details about the model are available elsewhere(7, 35). In summary, the model combines two cohorts (mothers and infants) and is split into three sections: pregnancy, childhood (birth to 15 years) and lifetime (100 years maximum). In each section smoking-related morbidity and mortality is applied to both cohorts to account for the effects of smoking. Costs and QALYs are accumulated in all sections for both cohorts.

**Figure 1:**
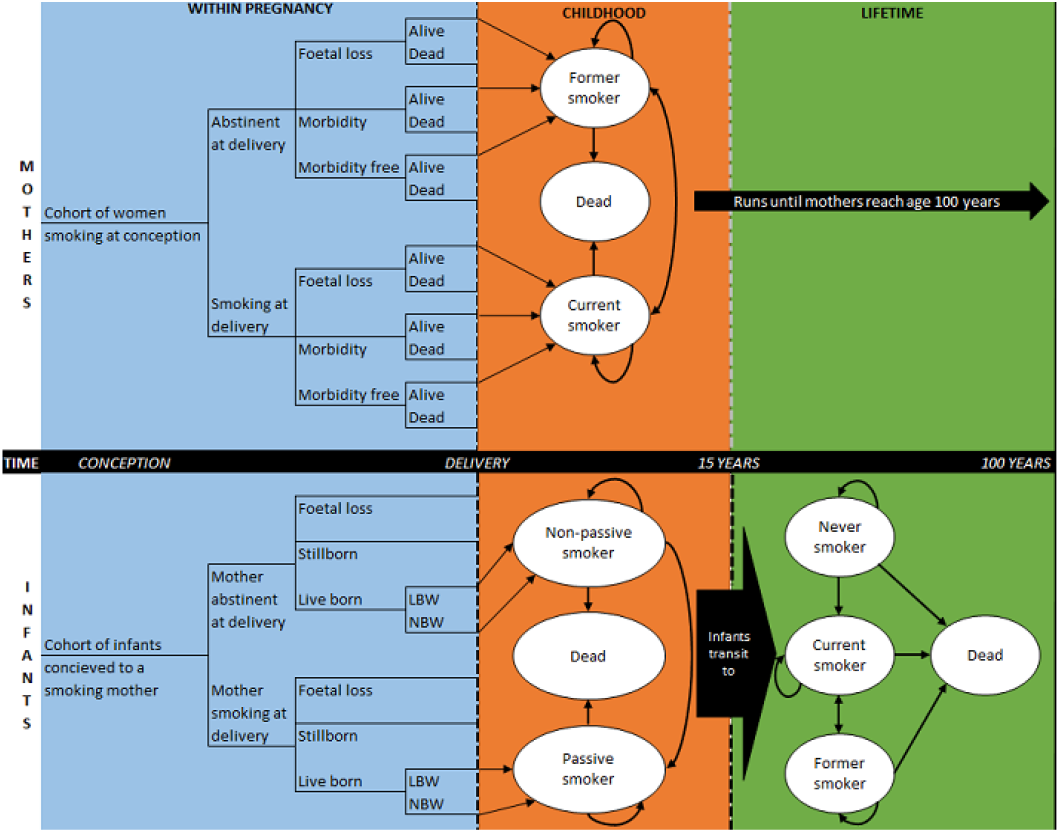
Model structure (Jones et al) (35)

#### Parameters

Treatment costs and quit rates from the CPIT III trial were input into the model for base-case and sensitivity analyses (Table 2), further details of existing parameters in the model are available elsewhere(7). Similar to the short-term analysis, late-pregnancy was used as a proxy for birth.

**Table 2:** ESIP model input parameters.

| Parameter | Input value | Source |
| --- | --- | --- |
| **Base case** |  |  |
| Cohort size | 944 | CPIT III trial data |
| Year of pregnancy | 2019 | CPIT III trial data |
| Age of mother | 28 | Mean age of participants in CPIT III (age at baseline) |
| Discount rate for costs/QALYs | 3.5% | NICE reference case(13) |
| Quit rates at late-pregnancy | Financial incentives 0.268 (SE 0.020)<br>Control 0.123 (SE 0.015) | CPIT III trial data (quit rate at late-pregnancy) |
| Cost of intervention and comparator | Financial incentives £262 (SE 11.7)<br>Control £91 (SE 6.40) | CPIT III trial data |
| Willingness to pay threshold | £20,000 | NICE reference case (13) |
| **Sensitivity analyses** |  |  |
| 1a - Gaming: quit rates | Financial incentives 0.239 (SE 0.020)<br>Control 0.098 (SE 0.015) | CPIT III trial data (quit rate at late-pregnancy – adjusted for within trial |
|  |  | gaming – incentive<br>reduce by 11%, control<br>reduce by 20%) |
| 1b - Gaming: quit rates | Financial incentives 0.239 (0.020)<br>Control 0.109 (0.015) | CPIT III trial data (quit rate at late-pregnancy – adjusted for within trial gaming –reduce incentives and control quit rates by 11%) |
| 2 - Self-report quit rate (late-pregnancy) | Financial incentives arm 0.359 (SE 0.022) Control arm 0.185 (SE 0.018) | CPIT III trial data (self-reported quit rate, with missing smoking status replaced with smoker – as per Russell standard) |
| 3a - Reduce incentive amount (max £160) and lower quit rate: Too, et al. (14) | Cost of intervention £131 (SE 13.265)<br>Quit rates incentives arm 0.046 (SE 0.005) and control arm 0.025 (SE 0.003) | Too et al., based on £160 maximum amount of vouchers and applying 24 weeks quit rate as proxy for late-pregnancy |
| 3b - Increasing incentive amount (max £800) and improved quit rate: Lussier et al. (35) | Cost of intervention with £800 maximum vouchers £463 (SE 47.4)<br>Quit rate for incentive arm 0.34 (SE 0.035) | Based on Lussier et al paper £800 maximum incentives and quit rate of 34% |
| 4 - Relapse rate in ESIP model replaced with CPIT III trial data | CPIT III 1-year relapse rate based on 6 months post-partum and extrapolated 0.814 (SE 0.079)<br>Model original 1-year post- | CPIT III trial data – 1 year relapse rate in M Jones model is taken from M Jones |
|  | partum relapse rate 0.47 (SE 0.046) | systematic review, in this sensitivity analysis it is replaced with CPIT III relapse rate for 6 months post-partum which is extrapolated to 1 year |

#### Analysis

Probabilistic sensitivity analysis (PSA) was conducted to allow characterisation of uncertainty in parameters. Base-case results for the following scenarios are presented: mother (pregnancy and lifetime), infant (pregnancy, childhood and adulthood) and combined lifetime (mother and infant).

Six sensitivity analyses explore the effect of varying model input parameters on results: 1) gaming (a) using quit rate based on gaming in trial (separate arms) and (b) based on gaming in incentives arm; 2) self-reported late-pregnancy CPIT III quit rate; 3) varying incentive amount and possible impact on quit rates based on (a) Too et al. findings(14); decreased incentive amount and lower quit rate and (b) based on Lussier et al. findings(36); increased incentive amount and improved quit rate, and 4) applying CPIT III 6-months post-partum quit rate, extrapolated to one-year, replacing existing one-year relapse parameter in the ESIP model.

Results are presented as mean and incremental cost and outcome for each arm with 95%CI. ICERs are presented with a 95%CI and probability of cost-effectiveness at £20,000 willingness-to-pay threshold (where appropriate).

## RESULTS

### Within-trial analysis

944 participants were randomised, three withdrew and asked that their data be removed, these participants were excluded from the economic evaluation in line with the primary outcome analysis. 941 participants remained in the analysis, 471 in the financial incentive arm and 470 in the control arm.

#### Missing data

Amount of missing data was similar in each arm; total costs 12% in both arms, late-pregnancy QALYs 21% and 25% for incentives and control respectively, post-partum QALYs 38% and 40% for incentives and control respectively (Table 3). 330 participants were missing either total costs or late-pregnancy QALYs, leaving 611 complete cases.

**Table 3:** Missing data.

| Variable | Incentives arm<br>(n=471)<br>N (%) | Control arm<br>(n=470)<br>N (%) | Total (n=941)<br>N (%) |
| --- | --- | --- | --- |
| Stop smoking services costs | 43 (9%) | 49 (10%) | 92(10%) |
| Nicotine replacement therapy costs | 56 (12%) | 57 (12%) | 113 (12%) |
| Total costs | 56 (12%) | 57 (12%) | 113 (12%) |
| Quitters – late-pregnancy | 59 (13%) | 39 (8%) | 98 (10%) |
| Quitters – post-partum | 94 (20%) | 92 (20%) | 186 (20%) |
| Utilities – baseline | 1 (0.2%) | 1 (0.2%) | 2 (0.002%) |
| Utilities – late-pregnancy | 101 (21%) | 115 (24%) | 216 (23%) |
| Utilities – post-partum | 133 (28%) | 132 (28%) | 265 (28%) |
| Quality-adjusted life years – late-pregnancy | 101 (21%) | 116 (25%) | 217 (23%) |
| Quality-adjusted life years – post-partum | 177 (38%) | 186 (40%) | 363 (39%) |
| Missing late-pregnancy QALYs or costs | 157 (33%) | 173 (37%) | 330 (35%) |

#### Number of vouchers issued

337 vouchers (71.4% participants in incentives arm) were issued for initial engagement with services and setting a quit date, 171 (36.2%) were issued at 4-week quit stage, 138 (29.2%) at 12-week quit stage and 150 (31.8%) at late-pregnancy. Three vouchers were either stolen or sent and not received, one each for engagement and setting quit date, four-week quit stage and late-pregnancy quit, these were excluded from the analysis. 344 (72.9%) participants in the incentives arm received one or more vouchers.

#### Base-case analysis

The incentives arm was more costly than control in the short-term, this difference was driven by neonatal costs (£,1723 v £982 incentives and control arms respectively) (Table 4). Intervention costs are higher in the incentives arm compared to control (£268 v £91), with £152 of those costs relating to vouchers.

**Table 4:** Short-term costs breakdown (unadjusted)

|  | **Incentives**<br>**Mean (SE)** | **Control**<br>**Mean (SE)** |
| --- | --- | --- |
| **Issued vouchers & postage** | £152 (7.63) | £0 (N/A) |
| **Stop smoking services** | £52 (2.54) | £41 (2.30) |
| **Nicotine replacement therapy** | £64 (5.15) | £50 (4.43) |
| **Total intervention costs** | *£268 (11.9)* | *£91 (6.40)* |
| **Neonatal** | £1,723 (548) | £982 (378) |
| **Total unadjusted costs** | £1,991 | £1,073 |

Adjusted results show total costs in the incentives arm are £637 (95%CI -£872 to £2,160) higher than in the control arm (Table 5). Late-pregnancy quit rate was higher in the incentives arm compared to control arm (0.268 v 0.123), a difference of 0.144 (95%CI 0.094 to 0.194). The incremental cost per late-pregnancy quitter was £4,400. QALYs were slightly higher in the incentives arm compared to the control arm (0.339 v. 0.335), a difference of only 0.004 and a 95%CI which crosses zero, (95%CI -0.163 to 0.175), the incremental cost per QALY is £150,000, which would not be considered cost-effective given the UK willingness-to-pay threshold of £20,000/QALY(13).

**Table 5:** Short-term analysis results.

| **Analysis** | **Cost/effect** | **Incentives**<br>**Mean**<br>**(Standard**<br>**error)** | **Control**<br>**Mean**<br>**(Standard**<br>**error)** | **Incremental** | **Incremental**<br>**Cost-**<br>**effectiveness**<br>**Ratio** |
| --- | --- | --- | --- | --- | --- |
| **Base-case**<br>**(cost per late-**<br>**pregnancy**<br>**quitter)** | **Cost** | £1,799 (21) | £1,161<br>(14) | £637 (-£872 to<br>£2,160) |  |
|  | **Quitter** | 0.268<br>(0.020) | 0.123<br>(0.015) | 0.144 (0.094 to<br>0.194) | £4,400 per<br>quitter |
|  | **QALY** | 0.339<br>(0.002) | 0.334<br>(0.002) | 0.004 (-0.143<br>to 0.150) | £150,000 per<br>QALY |
| **S1: including**<br>**miscarriage**<br>**n=18/944** | **Cost** | £1,805<br>(153) | £1,154<br>(98) | £652 (-£934 to<br>£2,245) | £151,000 per<br>QALY |
|  | **QALY** | 0.339 (0.01) | 0.335<br>(0.01) | 0.004 (-0.138<br>to 0.156) |  |
| **S2: self-reported quit – late-pregnancy missing n=58 replaced as smoker status** | **Cost** | £1,799 (21) | £1,161 (14) | £637 (-£872 to £2,160) | £3,700 per quitter |
|  | **Quitter** | 0.359 (0.022) | 0.185 (0.018) | 0.174 (0.118 to 0.230) |  |
| **S3: gaming - 11% incentives and 20% controls** | **Cost** | £1,799 (21) | £1,161 (14) | £637 (-£872 to £2,160) | £4,500 per quitter |
|  | **Quitter** | 0.239 | 0.098 | 0.141 |  |
| **S4: gaming - more conservative estimate 11% in both arms** | **Cost** | £1,799 (21) | £1,161 (14) | £637 (-£872 to £2,160) | £4,900 per quitter |
|  | **Quitter** | 0.239 | 0.109 | 0.130 |  |
| **S5: post-partum quitter** | **Cost** | £1,799 (21) | £1,161 (14) | £637 (-£872 to £2,160) | £24,000 per quitter |
|  | **Quitter** | 0.079 (0.01) | 0.051 (0.01) | 0.027 (-0.004 to 0.059) |  |
| **S6: post-partum QALY** | **Cost** | £1,799 (21) | £1,161 (14) | £637 (-£872 to £2,160) | £106,000 per QALY |
|  | **QALY** | 0.405 (0.01) | 0.399 (0.01) | 0.006 (-0.126 to 0.141) |  |
| **S7: complete case (n=611)** | **Cost** | 1,821 (159) | 1,589 (139) | £232 (-1,718 to 2,102) | £40,000 per QALY |
|  | **QALY** | 0.339 (0.02) | 0.333 (0.01) | 0.006 (-0.171 to 0.199) |  |
| **S8: postage £6** | **Cost** | £1,804 (21) | £1,161 (14) | £644 (-£748 to £2,432) |  |
|  | **Quitter** | 0.268<br>(0.020) | 0.123<br>(0.015) | 0.144 (0.094 to<br>0.194) | £4,500 per<br>quitter |
|  | **QALY** | 0.339<br>(0.002) | 0.334<br>(0.002) | 0.004 (-0.143<br>to 0.150) | £151,000 per<br>QALY |
| **S9: postage**<br>**£0** | **Cost** | £1,793 (21) | £1,162<br>(14) | £631 (-£722 to<br>£2,265) |  |
|  | **Quitter** | 0.268<br>(0.020) | 0.123<br>(0.015) | 0.144 (0.094 to<br>0.194) | £4,400 per<br>quitter |
|  | **QALY** | 0.339<br>(0.002) | 0.334<br>(0.002) | 0.004 (-0.143<br>to 0.150) | £158,000 per<br>QALY |

\*GLM regression results; QALY-quality adjusted life-year; S-sensitivity analysis

Uncertainty is illustrated on the cost-effectiveness plane and CEAC (Figure 2). Bootstrapped samples cover all four quadrants of the cost-effectiveness plane showing uncertainty in both cost and QALY results. There is less uncertainty in costs; most samples indicate higher costs in the incentives arm compared to control (above horizontal axis). At £20,000 willingness-to-pay threshold the CEAC shows a 36% chance of incentives being cost-effective in the short-term, increasing to 40% at £30,000, and not going above 47% up to £120,000 willingness-to-pay. Incentives are unlikely to be considered cost-effective in the short-term given the uncertainty regarding QALY gains.

**Figure 2:**
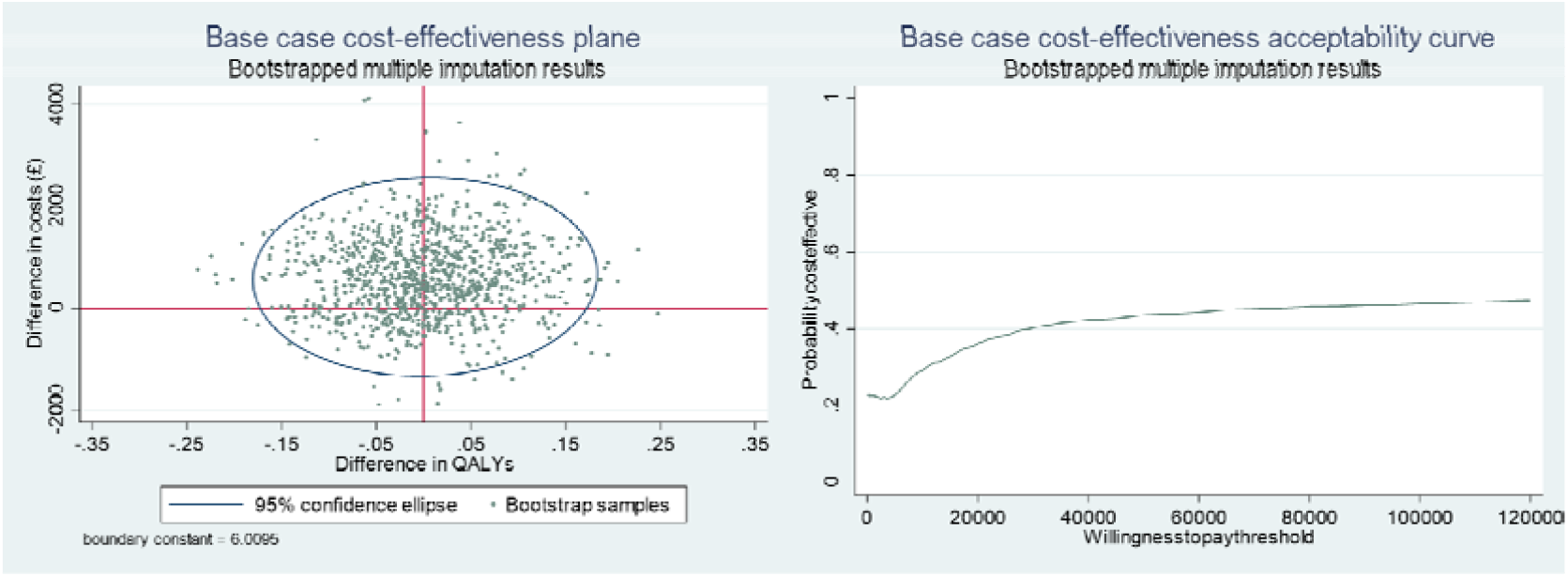
Base-case cost-effectiveness plane and cost-effectiveness acceptability curve.

#### Sensitivity analyses

Results confirm base-case results with cost per quitter ranging from £3,700 (self-reported quit rate) to £24,000 (post-partum quit). Cost per QALY results range from £40,000 (complete case) to £158,000 (£0 postage) (Table 5).

### Lifetime analysis

#### Base-case results

Results show that incentives would be considered highly cost-effective, given the UK willingness-to-pay threshold of £20,000, for mother lifetime, infant end of childhood, infant adulthood and combined mother and infant (lifetime) scenarios (Table 6). For ‘maternal end of pregnancy’ scenario, the probability of being cost-effective is 0%, with an incremental cost per QALY of £44,427. This is a similar conclusion to short-term results and unsurprising given health benefits of quitting smoking are not immediate. Combined lifetime mother and infant results estimate cost savings of £37 (95%CI -£35 to £106) and QALY gains of 0.171 (95%CI 0.124 to 0.229). These results show that introducing financial incentives to usual care is a dominant strategy (cost saving and QALY gaining) using a lifetime horizon. The probability of being cost-effective is 100%.

**Table 6:**
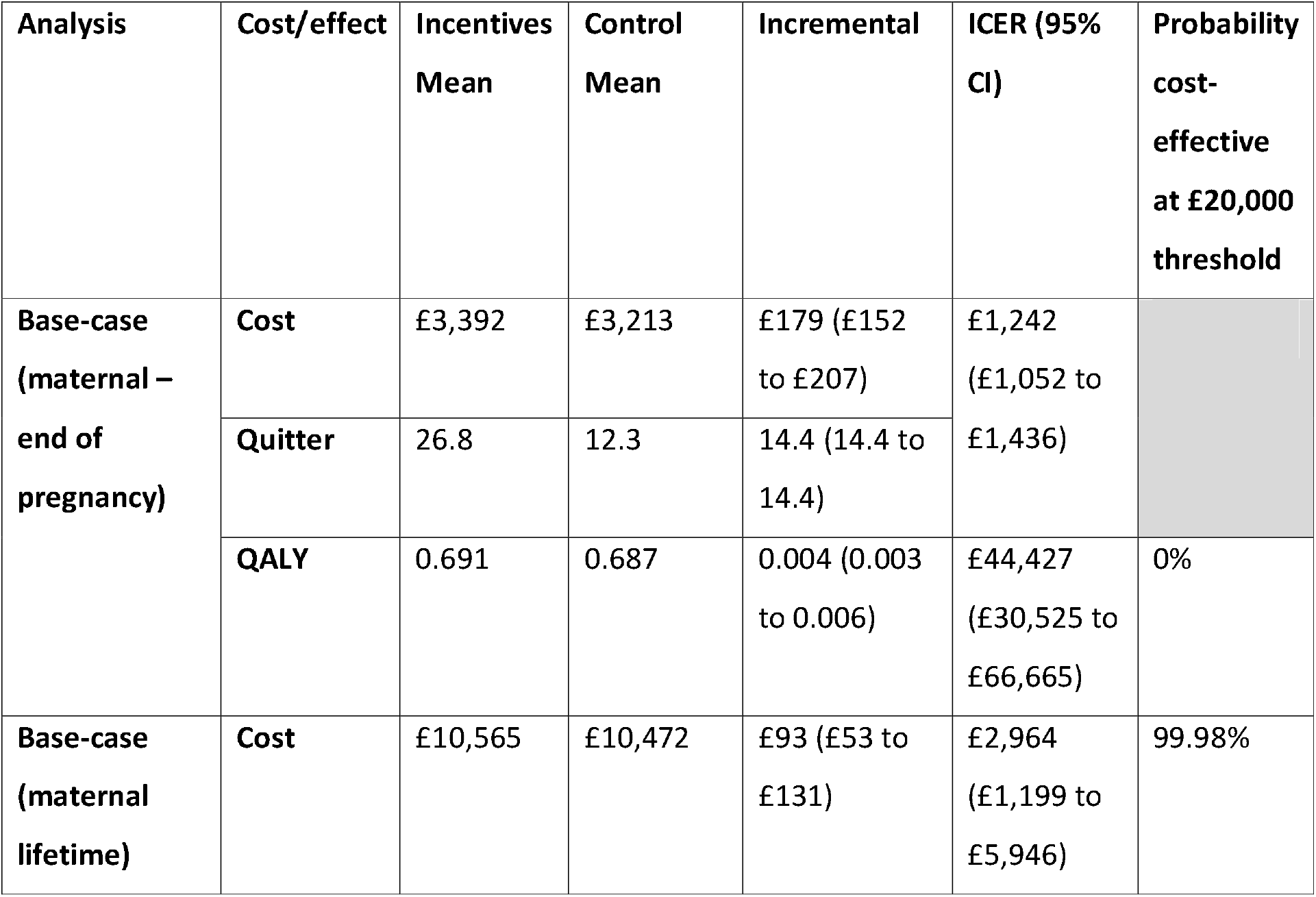
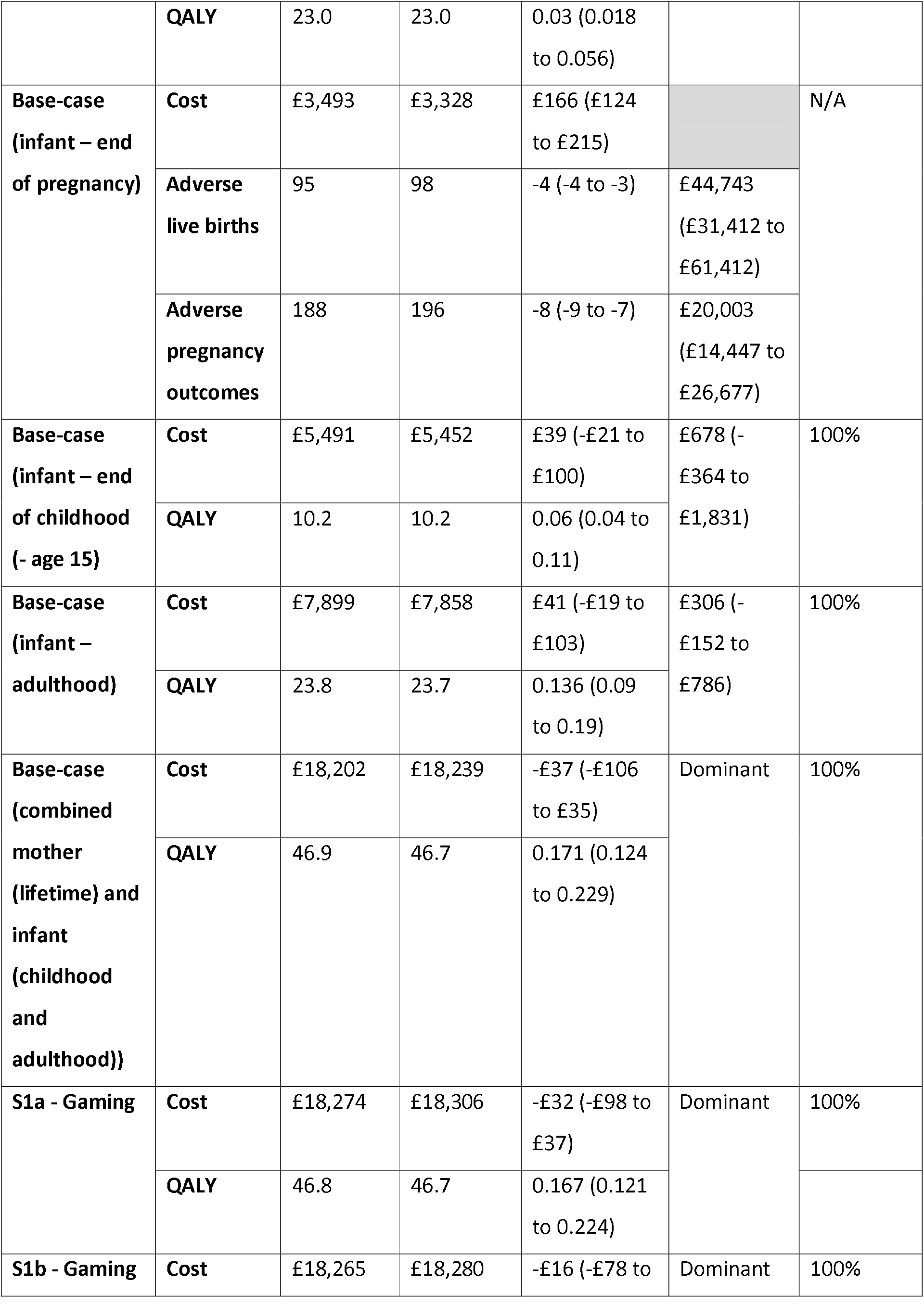
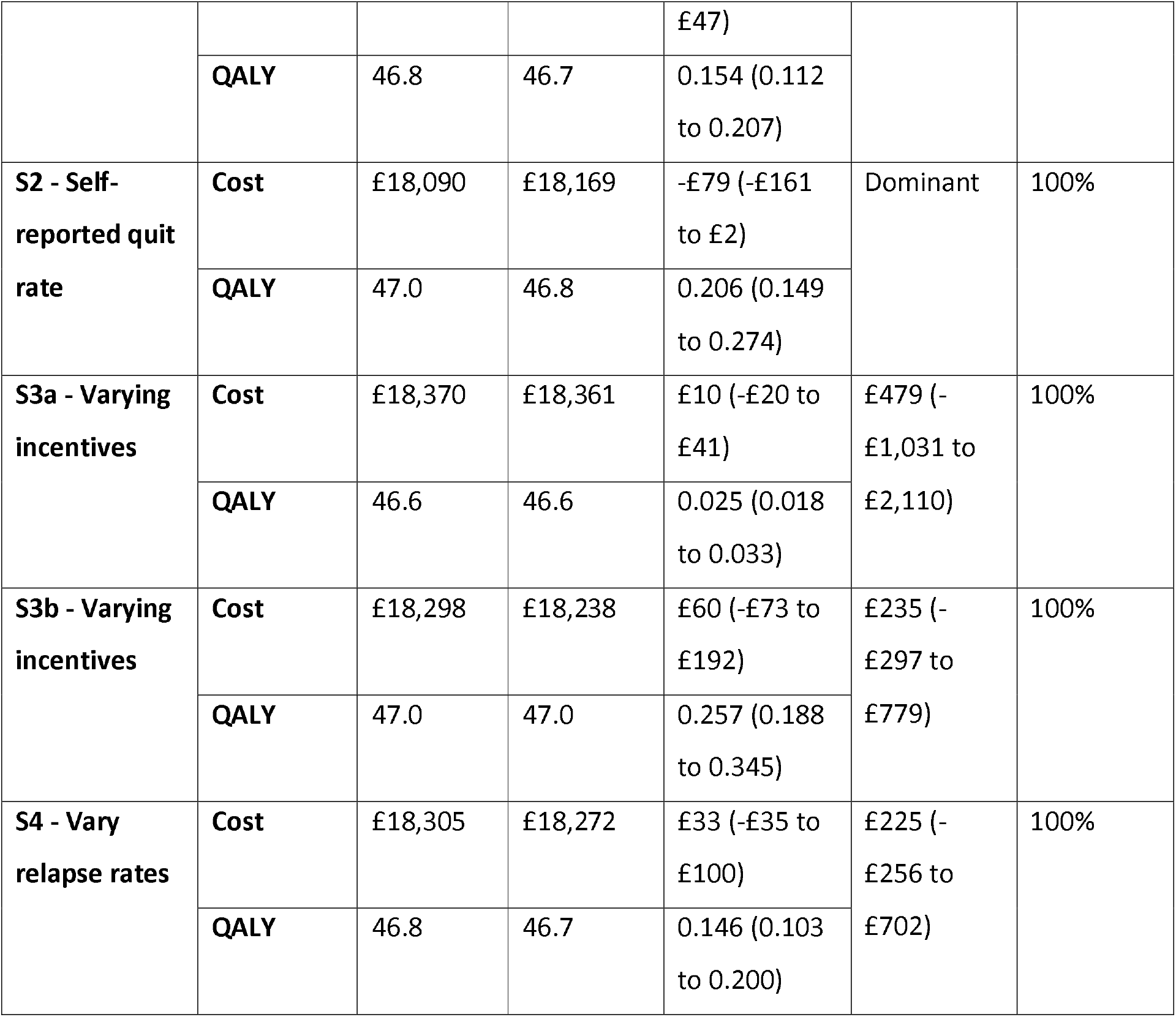
Lifetime analysis results.

Combined mother and infant (lifetime) results indicate little uncertainty (Figure 3). 10,000 PSA samples show higher QALYs in the incentives arm compared to control in all samples and lower cost in most samples, where incentives are dominant (cost saving and QALY gaining).

**Figure 3:**
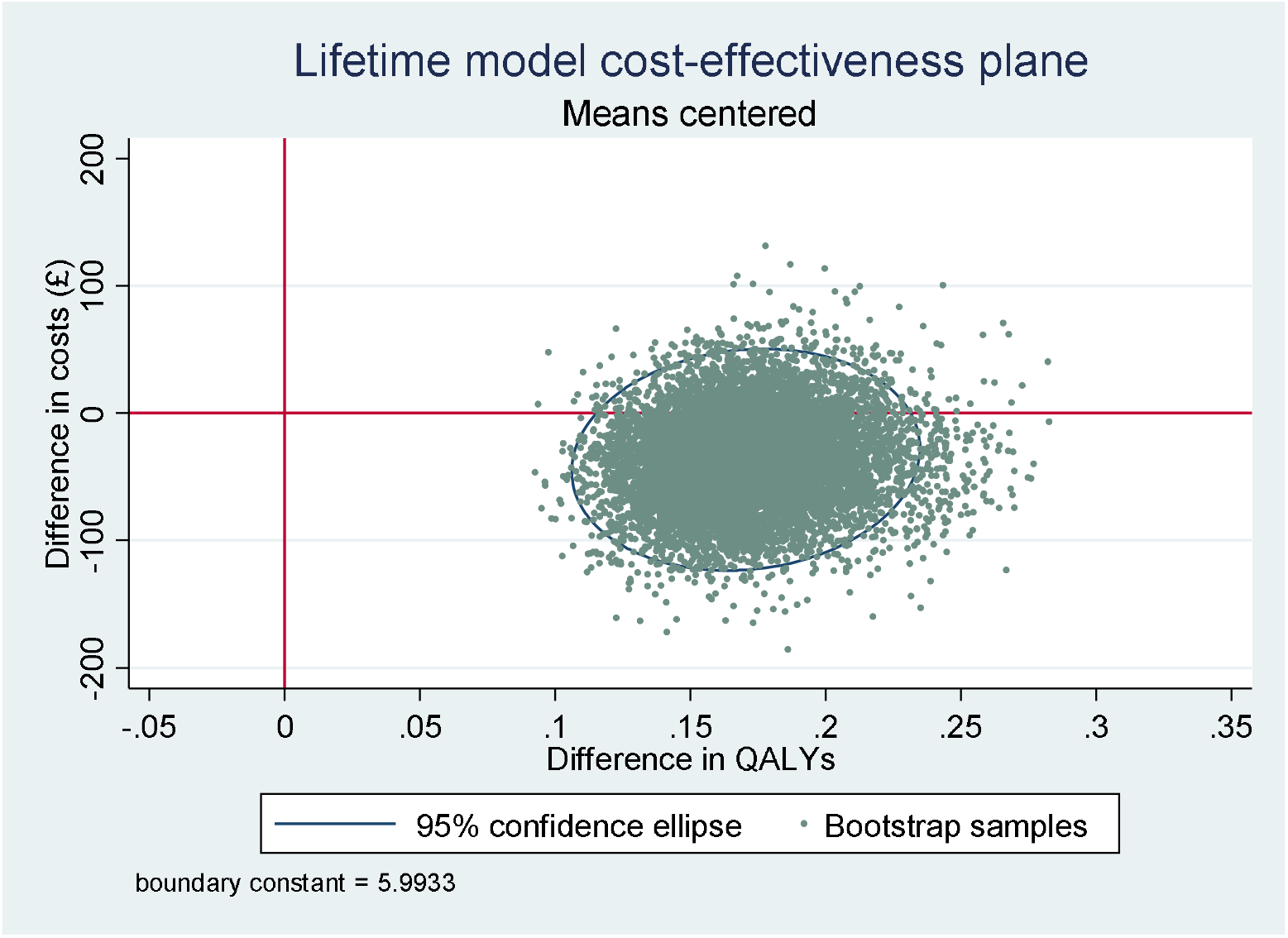
Lifetime model cost-effectiveness plane.

#### Sensitivity analyses (mother and infant lifetime)

Results confirm base-case long-term results; financial incentives would be considered highly cost-effective. Incremental cost per QALY ranges from a dominant strategy for self-reported quit rate and both gaming scenarios to an ICER of £479 (95%CI -£1,031 to £2,110) for using Too et al (14).

## DISCUSSION

Short-term cost per quitter was £4,400 and cost per QALY was £150,000; there was little uncertainty in difference in quit rates between arms, however differences in costs and QALYs were inconclusive. The long-term analysis found offering incentives to be cost saving (£37) with improved health benefits (quitters and QALYs). Over a lifetime offering financial incentives in addition to usual care is highly cost-effective for mother and infant compared to usual care only. Sensitivity analyses for the short- and long-term time horizons confirmed these results.

The CPIT II trial reported short-term cost per late-pregnancy quitter of £1,127 (12), lower than the present trial cost per quitter of £4,400, this difference is largely due to the inclusion of neonatal costs in the present analysis. CPIT II did not report a short-term cost per QALY but reported a model-based lifetime cost per QALY of £482. This is lower than the £2,964 reported in this present study for a maternal lifetime cost per QALY. An evaluation of the implementation of maximum £160 financial incentives in NHSGG&C reported cost per quitter at four- and 12-weeks of £517 and £546 respectively (14), again lower than the present study short-term cost per quitter but restricted to intervention costs and shorter time frame. A study assessing the cost-effectiveness of up to $500 financial incentives for mothers receiving Medicaid reported a cost per 6-months post-partum quitter of $3,399(37). Finally, a recent study assessing the cost-effectiveness of offering maximum $1,225 incentives reported an ICER of $23,511 per QALY at 24-weeks post-partum(15). However, neither of these two latter studies report lifetime cost per quitter or QALY.

During the trial three duplicate vouchers were issued and treated as a research cost. If financial incentives were implemented in a healthcare setting it is likely that duplicate vouchers would occur at additional cost, as well as postage and staff time administering financial incentives in a real-world setting. Scenario analyses explored alternative postage costs, with minimal impact on ICER results. In terms of implementation, it would be appropriate to consider alternative voucher types and distribution methods to improve efficiency, such as electronic vouchers or mobile phone app which could be ‘topped up’ remotely or by way of codes when self-reported quits are validated. These would be relatively cheap, compared to the trial methods employed for voucher distribution.

Strengths of the economic evaluation include the trial being pragmatic, reflecting actual practice at seven sites across Scotland, Northern Ireland and England and providing evidence on the success of financial incentives in real-world situations. Research shows maternal smoking during and after pregnancy can have serious negative consequences on the infant. The effects of maternal smoking were incorporated in our long-term analysis to reflect the impact on mothers and infants. There is little evidence on quit rates and quality of life post-partum, however we collected these outcomes six-months post-partum, although this data was subject to missingness it provides additional evidence in this field. Further, we input CPIT III post-partum relapse rates into the ESIP model to reflect the rates witnessed in a trial, the resulting ICER showed results would be considered cost-effective. SSS support varied between sites and data on individual NRT and SSS was limited to five of the seven sites due to challenges during the Covid-19 pandemic. Costs for the two additional sites were based on data collected in the trial database, potentially reducing precision of our analyses. CPIT III trial did not collect neonatal stay data, therefore prematurity status and severity were used as a proxy indicator for neonatal stays.

There is little and variable evidence on whether the amount of financial incentives offered impacts quit rate. Previous research suggests increasing incentive amount improves the effectiveness in substance use and smoking cessation(36, 38). However more recent research has shown no clear evidence of this link in health behaviour change(10, 39, 40), and indeed with increasing incentive amounts, diminishing returns would set in due to a cubic trend(40). Another barrier to estimating links between amount of incentive and magnitude of effect is that the success of financial incentives is also dependent on the level of cessation support (41); in CPIT III cessation support was found to vary by site. Further research is needed into varying the amount of incentive and the resulting impact on effectiveness.

## Conclusions

Offering financial incentives alongside usual care is effective at improving quit rates in the short-term and is highly cost-effective for mother and infant over a lifetime. This research should prompt healthcare providers to offer financial incentives, alongside usual care, to pregnant women who smoke to encourage engagement with support service and improved quit rates.

## Data Availability

All requests for data should be submitted to NM for consideration. Access to anonymised data may be granted following review by the Trial Management Group and agreement of NM and DT. The data are not publicly available due to privacy or ethical restrictions.

## Acknowledgements

We would like to acknowledge the wider CPIT team: Margaret McFadden, Sinead Watson, Alison Dick, Helen Tilbrook, Ada Keding, Judith Watson, Frank Kee, David Torgerson, Catherine Hewitt, Jennifer McKell, Pat Hoddinott, Fiona M Harris, Isabelle Uny, Michael Ussher. We would also like to acknowledge Matthew Jones for the use of the ESIP model and his help and support with its use.

## FOOTNOTE

There is a deviation from protocol as mean costs and outcomes are presented with standard errors not standard deviations as stated in the protocol(21).

## PATIENT AND PUBLIC INVOLVEMENT

No patient involvement.

## ETHICS AND DISSEMINATION

Ethics approval was received from NHS West of Scotland Research Ethics Committee 15th August 2017.

## AUTHORS’ CONTRIBUTIONS

N McMeekin: Methodology; formal data analysis; drafting manuscript; review and editing

L Sinclair: Data curation; investigation; project administration; review and editing

L Robinson-Smith: Data curation; investigation; project administration; review and editing

A Mitchell: Data curation; methodology; review and editing

L Bauld: Funding acquisition; review and editing

D Tappin: Funding acquisition; review and editing

K.A. Boyd: Conceptualisation; funding acquisition; methodology; supervision; review and editing

